# Community Needs Assessments in a Student-Run Clinic serving a West Philadelphia Neighborhood

**DOI:** 10.1101/2020.04.15.20066563

**Authors:** Daniel J. Arenas, Dania D. Hallak, Rommell Noche, Gilberto Vila-Arroyo, Swathi Raman, Arthur Thomas, Sara Zhou, Adriana Richmond, Raisa Rauf, Irene Su, Sourik Beltrán, Tim Lee, Rebecca Abelman, Casey Baginski, Heather A. Klusaritz

## Abstract

**Background:** While Community Needs Assessments (CNAs) are an important tool for Student-Run Clinics (SRCs) to understand local communities’ healthcare needs, few studies have evaluated CNAs and their impact on care provided at SRCs.

**Objective:** Evaluate results from two CNAs of an SRC in East Parkside, Philadelphia to better comprehend (1) community awareness and opinions regarding the SRC and (2) local healthcare concerns and access.

**Methods:** 58 and 105 East Parkside residents were surveyed in 2011 and 2015 respectively. The results were analyzed to quantify various health-related measures in the community.

**Results:** Results showed high rates of hypertension, asthma, and diabetes. Rates of pap-smear and hypertension screening matched national averages while mammograms and colonoscopies were below national rates. Both CNAs showed that less than 40% of community members were aware of the clinic’s existence.

**Conclusions:** CNAs can provide valuable insights regarding local health needs which can inform future healthcare interventions.

---

Community Needs Assessments (CNAs) are an important tool that allow healthcare institutions to assess and improve the quality of care being provided to a particular community. Methodologies to conduct CNAs vary from performing in-clinic patient surveys,^1^ reviewing existing data from nearby hospitals or community service partners,^2^ reviewing published studies,^3^ to hosting or attending town halls and focus groups.^4,5^

Student-Run Clinics (SRCs) play a crucial role in the healthcare safety net as well as provide valuable community-engaged training opportunities.^6–10^ SRCs are beneficial to both students and patients in that students receive interdisciplinary, community health experience, while providing healthcare to often vulnerable communities in such a way that yields high patient satisfaction and significant economic and health impacts.^11,12^ Similar to other innovative health care interventions serving vulnerable communities, resource allocation is an important consideration for SRCs as they must identify and modify the interventions that would maximize their impact on the community. Therefore, CNAs can be an important tool for SRCs and other health providers working to alleviate healthcare disparities. There is already evidence that patient-focused CNA studies have been useful in improving care quality as well as identifying local health disparities.^13–16^ CNAs could therefore be utilized as a tool to eliminate disparities in both access to healthcare and resource utilization.^17–19^ Accordingly, CNAs have become increasingly common as many government agencies now offer tax incentives for hospitals that make efforts to assess their local communities’ health needs.^20^

Despite the benefits of CNAs for SRCs, there has been little uptake in their use. A recent systematic review conducted by the authors showed that, as of 2017, there were less than three published studies that directly surveyed patient populations by going into the communities served by SRCs and other free clinics.^21^ The review demonstrated an overall rise in SRCs use of CNAs.^4,5^ Nevertheless, few studies described the role of CNAs in well-established clinics and similarly few publications performed the CNAs using quantitative methods. The review suggested a need for more free, student-run clinics to perform CNAs as CNAs are a useful tool in identifying local health needs,^22–24^ diagnosing gaps in social determinants of health,^25^ improving health education,^26^ and identifying community concerns.^22–28^

This study addresses the gap in the CNA literature by critically evaluating the methodology and results of two CNAs performed by the United Community Clinic (UCC), an SRC that serves a population in the East Parkside neighborhood of West Philadelphia, PA. The CNAs were shown to be an efficacious way to assess the clinic’s target population healthcare accessibility and to survey the health concerns patients had both for themselves and their community. For the first time in the literature, this study shows that CNAs can also be a useful tool for evaluating “brand awareness” of SRCs, or the percentage of community members who are aware of the clinic’s existence. The authors also comment on the lessons learned from the CNAs, as well as the limitations faced while analyzing data from CNAs performed by different student generations.

## Methods

### Description of United Community Clinic

UCC is a general primary clinic based out of the First African Presbyterian Church in the East Parkside community of West Philadelphia. The clinic provides a wide range of free medical services with a particular focus on preventative medicine. The majority of patient encounters at UCC involve physical exams necessary for job applications, driving licenses, and sport evaluations. The clinic runs from 6PM to 9PM every Monday night and is staffed by undergraduate, medical, nursing, dental, social, podiatry, and optometry students, as well as faculty advisors consisting of interns, residents, nurse practitioners, and attendingphysicians. All volunteers are associated with the University of Pennsylvania.

### Survey Overview

First in 2011 and then in 2015, two distinct generations of student leaders, volunteers, community members, and advisory boards designed CNAs targeted towards the East Parkside community of west Philadelphia. They designed the process to align with the Public Health Management Corporation (PHMC) Household Health Survey (HHS) domains. S1 of the supplementary documents provides the original surveys for each CNA. The survey was divided into eight sections addressing participant demographics, clinic experience, health status, healthcare access, preventative care, insurance, and other general questions. The survey was verbally administered to each willing participant.

### Survey Administration

Undergraduate, nursing, medical, and social work students who volunteered at UCC were recruited to administer the survey. Students attended one mandatory training workshop to discuss survey administration techniques and how to standardize their delivery.

In the 2011 and 2015 CNAs, the target population was divided into fifteen blocks. One pair of students was assigned to each of the fifteen blocks and instructed to recruit participants from their respective block. The survey was administered on Saturday mornings. Students knocked on doors to recruit participants and informed potential participants of the opportunity to receive a gift card upon completion of the survey. Students then proceeded to administer surveys outside of the residents’ homes. For the 2015 CNA, the residents who completed the survey were given $10 ShopRite gift cards, funded by the student-clinic budget. Both CNAs were approved by University of Pennsylvania IRB as quality improvement projects for the SRC. No patient identifiable information was collected.

### Inclusion and Exclusion Criteria

The 2011 study interviewed 58 East Parkside residents, and the 2015 study interviewed 105. All participants of both studies were East Parkside residents who were surveyed at home during the survey administration hours after agreeing to be included in the study.

### Statistical Analysis

Confidence intervals (CI) for percentages were calculated using the Agrousti-Coull method to account for the randomness of the binomial distribution.^29^ All intervals presented denote a 95% CI. Significance in changes between percentages across years were evaluated using non-paired *t*-test of proportions with continuity correction.^30,31^ Two-tailed tests were performed for testing differences, and one-tailed tests were performed for comparisons having a-priori hypotheses of increase (i.e. increase in health insurance coverage after the Affordable Care Act). Comparison to national / regional averages were performed using one-sample *t*-tests. Statististical significance was set at a type I error of 0.05. Means and standard deviations (denoted by parentheses) are reported for all continuous variables. All statistical calculations were performed using R.

## Results

### Community Profile and Clinic Awareness

Table 1 shows the demographic information of the survey respondents for both the 2011 and 2015 CNAs. Over 95% of respondents in both CNAs confirmed being residents of East Parkside. Nearly half of the residents had lived in the community for more than 10 years, and over 89% of residents identified as black in both CNAs. The average ages for the 2011 and 2015 surveys were 44 (SD: 17) and 48 (SD: 17) years of age, respectively. About half of the respondents identified as male and the other half identified as female in both CNAs. In 2011, 37% [CI: 25-50%] of the interviewed community members reported awareness of the existence of UCC in their area, and 40% [CI: 31-50%] reported awareness in 2015. The change in clinic awareness was statistically non-significant (*p* = 0.85).

**Table 1.** East Parkside Demographics.

|  | **2011 CNA**<br>**(N=53)** | **2015 CNA**<br>**(N=104)** |
| --- | --- | --- |
| **Mean age, years (SD)** | 44 (17) | 48 (17) |
| **Female\*** | 56% | 53% |
| **Race** |  |  |
| Black | 89% | 92% |
| Hispanic | 6% | 1% |
| White | 2% | 3% |
| Other | 3% | 4% |
| **Parkside Residency** |  |  |
| <3 years | 33% | 18% |
| 3-10 years | 19% | 29% |
| >10 years | 48% | 53% |

\*For those who did not identify as male or female, the “other” option was included in the 2015 CNA, although nobody reported as such.

### Accessibility to Healthcare and Preventive Medicine

Health insurance accessibility was surveyed in both CNAs (Table 2). 76% [CI: 63–85%] and 88% ([CI: 80-93%], *p* = 0.039) of interviewees reported having health insurance in the 2011 and 2015 CNAs, respectively. The comparison of the two surveys is important as it correlates with the implementation of the Affordable Care Act starting in 2010.

**Table 2.** Health Insurance accessibility.

|  | **2011 CNA**<br>**(N=54)** | **2015 CNA**<br>**(N=102)** |
| --- | --- | --- |
| **Has Health Insurance** | 76% | 88% |
| **Type of Insurance** |  |  |
| Employer | 35% | 20% |
| Individual* | 2% | 14% |
| Medicaid | 37% | 33% |
| Medicare | 19% | 27% |
| Other | 2% | 5% |

\*includes insurance purchased individually through Health Exchange (ACA) in the 2015 CNA.

The 2015 CNA showed that about 20% [CI: 16-32%] of the population reported having been uninsured at some point in the previous 12 months. In both CNAs, over half of respondents reported having government-provided health insurance. Ninety percent of interviewees reported seeking health care when they were sick and 92% reported receiving regular preventive health care (Table 3). Forty-six percent of respondents (2011) and 47% (2015) had their preventive medicine provider in the neighborhood.

**Table 3.** Access to Healthcare.

|  | **2011 CNA**<br>**(N=54)** | **2015 CNA**<br>**(N=102)** |
| --- | --- | --- |
| **Do you receive care when sick?** |  |  |
| Yes | 93% | 91% |
| No | 7% | 9% |
| **Do you receive regular preventive care?** |  |  |
| Yes | 92% | 99% |
| No | 8% | 1% |
| **Is the place you go for care in your neighborhood?** |  |  |
| Yes | 46% | 47% |
| No | 54% | 53% |

The 2015 survey also provided information about adherence to preventive medicine guidelines. The highest participation rates were found in hypertension and pap-smear screenings; 89% [CI: 81-94%] reported having their blood pressure measured in the last year and 95% [CI: 88-98%] within the last 2 years. Ninety percent [CI: 78-96%] of women in the 21-65 age range reported having a pap smear in the last three years while 78% [62 – 88%] of women in the 40+ age range reported having a mammogram within the last two years. The lowest participation rates were found in colon cancer screenings, where approximately 46% [CI: 33-57%] of the community members in the 50-75 age range reported never having a colonoscopy or sigmoidoscopy. (Table 4). This self-reported adherence to colonoscopy screening guidelines was lower than the 67% state average.^32^ Similarly, only 50% [CI: 40-59%] of surveyed individuals reported seeing a dentist annually, a figure higher than the national average of 33%.^33^

**Table 4.** Preventive care. 2015 CNA results.

| **How long since last BP reading?** |  | **N = 98** |
| --- | --- | --- |
| 1 year ago or less |  | 89% |
| 1-2 years ago |  | 6% |
| >2 years ago |  | 5% |
| **How long since last Pap smear test? (Women ages 21-65)** |  | **N = 50** |
| 3 years ago or less |  | 90% |
| >3 years |  | 10% |
| **How long since last mammogram? (Women ages 40+)** |  | **N = 40** |
| 2 years ago or less |  | 78% |
| >2 years |  | 22% |
| **How long since last colonoscopy or sigmoidoscopy? (Ages 50-75)** |  | **N = 61** |
| 10 years ago or less |  | 49% |
| >10 years ago |  | 5% |
| Never |  | 46% |

### Health Status

Health status was surveyed in the 2015 CNA, but not the 2011 CNA. In 2015, 52% [CI: 41–62%] of the interviewees reported having been diagnosed with hypertension in their lifetime. This self-reported figure was significantly higher than the 28% prevalence of hypertension in the United States.^34,35^ Of those diagnosed with hypertension, 73% [CI: 59-84%] reported taking a prescribed medication. This number was significantly higher than the 25% adherence rate for Medicare and 45% rate for nationally.^36^ For diabetes, the prevalence was 20% [CI: 12-29%], and of those diagnosed, 75% [CI: 51-91%] were taking medication. Similarly, 23% [CI: 16-33%] of those surveyed had been diagnosed with asthma and 71% [CI: 49-87%] were taking a prescribed medication. Lastly, 31% [CI: 22-40%] of those surveyed reported fair or poor health while 50% [CI: 40-60%] reported overall poor health in the neighborhood.

## Discussion

Analysis of two CNAs performed by UCC shows that these assessments can yield helpful insights and statistics about the patient population’s health status, access to preventive medicine, and awareness of the clinic’s existence, all of which can be used to guide future clinic programming.

The CNAs found a higher than average prevalence of diabetes and asthma as well as access barriers to some preventive health resources including dental care, mental health services, and colonoscopies. Despite high rates of insurance, gaps in access existed, allowing for patient-centered expansion of clinic services. For example, given the high rates of hypertension found in the 2011 CNA, UCC developed a hypertension-focused clinic (Health Heart Bridge to Care), which focuses on long term hypertension management and facilitating linkage to care. Since the 2015 CNA found that there was also a need for diabetes resources in the target population, UCC established a new program that offers blood glucose testing to those at high risk for diabetes, programming to assist with long-term diabetes management and referrals, meetings with diabetes specialists, and workshops dedicated to healthy eating and increased activity for those who had diabetes or were at risk of having diabetes. In response to findings that indicated a large need for services such as colonoscopies and mammograms that could not be offered in-house by the SRC, UCC added an outside social work organization, Service Link, to aid in patient referral. Additionally, to address a high prevalence of asthma in the East Parkside community, UCC is currently working on providing inhalers for patients who need them.

An encouraging result was the statistically significant increase in community members who had health insurance from 78% in 2011 to 88% in 2015. The community may have been positively impacted by the implementation of the Affordable Care Act in 2010. However, the clinic must continue to monitor the situation as the future of the Affordable Care Act or an analogous program is unclear as of 2019.

Despite the founding of UCC in 1997, only 37% of interviewed community members in 2011 reported an awareness that the clinic existed. In 2015, there was only a statistically insignificant 3% increase in the awareness of the clinic. To our knowledge, this is the first quantitative report of clinic awareness in the literature. Since there are no reports in existing literature of brand awareness of SRCs, it cannot be deduced how 40% community awareness compares to other SRCs nationwide. The fact that awareness did not increase in 4 years does however suggest a need for active clinic promotion rather than simply relying on word of mouth.

In an effort to improve the 40% clinic awareness, UCC formed a special outreach committee in which UCC members attended community meetings, increased clinic advertising in popular community spaces such as churches. Increasing awareness of the clinic, even to those who do not need its services, has since become a top priority of UCC’s leadership. It is hoped that resultant improvements in clinic awareness will serve to further reduce the health disparities found in the East Parkside community.

The CNA also identified a need for services such as colonoscopies and mammograms that are not offered in-house at UCC. In response, UCC partnered with outside social work organization, Service Link, to aid in patient referral.

## Limitations

The greatest limitation of our research is that the CNAs were planned and performed by different student leaders, with a third group of students conducting a secondary analysis of the results. As the CNAs had been performed primarily for guiding clinic programming, the entire methodology of the 2011 CNA was not available for this study (for example, the exact map of the areas that were surveyed). The 2015 CNA worked to streamline methodology from the 2011 CNA, and therefore had a different questionnaire and survey strategy.

Several improvements could be made to the survey. Further questions about the community’s opinion of UCC, as well as the ways in which community members discovered UCC could have been beneficial in order to identify how UCC could reach more community members. Another important limitation of the CNA and of the statistical analysis that ensued was the ability to ensure random sampling. Due to logistics, our sample involved people who were at home on Saturday mornings introducing a factor of non-randomness.

Lastly, self-reported previous diagnoses of chronic diseases are not as reliable as cross-sectional studies of disease prevalence. All comparisons to national values must be taken in the context that self-reporting could add other confounding variables such as perception of health. Rather, these self-reported numbers were used as an indicator of the population’s concern as well as what preventive medicine interventions should be focused on.

An improvement on the 2011 survey design in the 2015 version was the inclusion of the gender option for those who identified as neither male nor female, although no respondents identified as such.

## Conclusions

CNAs performed by SRCs reveal information about demographics, health status, and health concerns of the communities that they serve. This information is crucial for evaluating and improving intervention and outreach efforts of SRCs. The CNAs performed in this study provide a sample methodology, as well as methodological critique, for other SRCs to incorporate CNAs into their community engagement. Future CNAs should attempt to elucidate more information about clinic awareness, ethnicity, and adherence to medication and preventive health guidelines.

## Data Availability

Primary data is presented in the manuscript tables

## Declarations

### Availability of supporting data

S1: Original survey for the 2015 CNA.

### Author’s contributions

HSK was part of the faculty leadership and led the two CNAs. Authors from the 2011 UCC generation, TL and RA, led the first CNA; RA, DH, and SR – authors from the 2015 UCC generation - conducted the second CNA. Authors from the 2017 UCC generation (RN, AT, IS, AR, RR, and DJA) analyzed the 2011 and 2015 CNAs. DJA led the analysis of the data, and along with SB, performed the statistical analysis. RN, DH, SR, AT, SZ and DJA wrote the first draft. All authors read consequent drafts, provided input, and approved final version.

## Acknowledgments

Special thanks to Ms. Jerri who welcomes patients and health providers with a smile and open arms to the church and clinic. AT and SB would like to thank the Perelman School of Medicine scholarship for support. DJA would like to thank the Gamble Scholarship.

